# Exploring Inequalities in Population Health: A Phenome-Wide Study Examining the Association between Education and 833 diseases in Denmark

**DOI:** 10.1101/2023.06.22.23291734

**Authors:** Anna Vera Jørring Pallesen, Jochen Mierau, Laust Hvas Mortensen

**Author notes:** **The corresponding author:** Anna Vera Jørring Pallesen.

## Abstract

**Objective:** The social processes that shape people’s environment and ways of living tend to create inequalities in health. Better education is often, but not always, associated with lower disease incidence and better prognosis through a multitude of different mechanisms. Most often, research in this area examine few diseases of interest, thereby creating an array of disjoint analyses that lack comparability. The objective of this study is to create a novel atlas covering 833 diseases illustrating the associated educational gradients across a wide spectrum of health outcomes.

**Method:** Longitudinal, register-based study conducted on all Danish residents aged 30 years and over (N=4,258,789) between 2012 and 2021. We applied a phenome-wide approach to assess associations between three ISCED-based educational levels (low, medium, high) and the incidence of 833 diseases defined by ICD-10 diagnostic codes. Age-standardized incidence rates were estimated using Poisson regression adjusted for sex and birth cohort.

**Results:** Educational inequalities were observed in virtually all diseases studied and were, thus, not limited to particular disease areas. We found incidence rates of the vast majority of diseases increased with higher educational level (N=569). For 77 diagnoses, we observed an inverse educational gradient. Lower educated individuals had, with few exceptions, the highest incidence rates for non-communicable and communicable diseases.

**Conclusions:** Our atlas provides a full and detailed representation of the association between education and disease incidence. It brings attention to all diseases – not only the most prevalent – and makes it possible to examine the role of education across the universe of diseases.

## Introduction

Population health is improving due to advancements in clinical diagnosis, medical treatment as well as societal development and prevention, however, the difference in health outcomes across groups of different socioeconomic position (SEP) is increasing (Vaupel et al., 2021; Dalton et al., 2008; van Raalte et al., 2018; Mackenbach et al., 2015). A study by van Raalte and colleagues (2018) showed that life expectancy had increased in all population groups in Finland from those of high through those of low SEP. But while life expectancy increased across all sub-populations, the widening health gap between individuals with high and low SEP remains an important public health issue (Marmot et al., 2020; Maldi et al., 2019).

Inequalities in population health are a result of social stratification, which leads to different economic, social and cultural advantages for different groups of people (Galobardes et al., 2007).These advantages are shown to be associated with better health outcomes and lower mortality in a substantial number of studies (Mackenbach et al., 2008; Pallesen et al., 2021; Torssander et al., 2016; Fonseca et al., 2020; Olsen et al., 2019; Pathirana and Jackson, 2018). However, the link between social stratifications and different health states are various and not unequivocal. Research has shown that the strength of this association varies depending on specific diseases under study whereas social advantages are not always related to better health (Davey Smith, 2013). For instance, skin, breast and prostate cancers are, often found to be more frequently diagnosed among persons who completed a higher education than persons with a lower educational attainment (Dalton et al., 2019). SEP may be defined in various ways such as income, occupation or education. They reflect different and correlated dimensions of socioeconomic position which may affect health outcomes similarly or differently depending on the outcome studied (Duan et al. 2022; Geyer et al., 2006). To provide a first exploration, we focus only on education in this paper but encourage future authors to focus on other dimensions of SEP (see also the Discussion).A large body of research explored how education acts as an important determinant of health. At the individual level, education leads to lasting cognitive changes, which enhances the ability for abstract thinking, sense of personal control, and motivation to solve problems and plan for the future (Kandel, 2007; van der Pol, 2011; Skeide et al., 2017; Heckman, Humphries, and Veramendi, 2017). Consequently, more educated individuals are better at identifying health risks and adapt their behaviour to minimise exposure to health hazards and pursue better health (Baker, 2014). On a structural level, education is linked to future economic circumstances and work environment, which too impacts health (Diderichsen et al., 2012).

The complex interplay between education and health, however, is often studied for one or few specific diseases at a time. In addition, the study populations differ widely from study to study. This creates an array of disjoint analyses that lack comparability. In this paper, we argue that examining an extensive list of diseases across a wide spectrum of morbidities will provide a fuller and more detailed representation of the health gap between individuals with high and low SEP. Applying the approach of the phenome-wide association studies (PheWAS), the objective is to create an atlas covering over more than 800 diseases and illustrating the associated educational gradients of each disease.

## Methods and material

For the present study we extend the approach of phenome-wide association studies (PheWAS) to study the role of a socioeconomic indicator (i.e., education) on the universe of diseases. This approach originated in genetics research to test for associations between one specific or combinations of more genetic variants and a wide selection of phenotypes in large populations (Hebbring, 2013; Bastarache et al., 2022). The method usually tests the genetic contribution for a specific phenotypic outcome. In the context of educational inequalities in health, diseases constitute a phenotypic outcome. Thus, the present study constitutes a compilation of longitudinal cohort studies each with a specific disease as outcome that are presented jointly to provide an atlas of health inequalities across the universe of diseases.

This study was conducted based on data from the national Danish registries. Specifically, we used data from the Danish Lifelines, the Danish National Patient Registry (DNPR) and the Education Registry. The Danish Lifelines describes the population in demographic terms such as reason and date of entering (e.g. birth, immigration) and leaving (e.g. death, emigration) the population. The Danish Lifelines, furthermore, draw on information from the Civil Registry, which was implemented in 1968 and contains basic information (e.g. sex, birthday) on all individuals. The DNPR contains information on hospital admissions. Since 1977, the registry has held information on all somatic hospital admissions, and from 1995 ambulatory and emergency department contacts were included as well as admissions to psychiatric departments. From 2003 it, additionally, became compulsory for private hospitals to report all patient contacts to the DNPR (Schmidt et al., 2015). Hence, the DNPR covered all inpatient and outpatient contacts at public and private hospitals in Denmark during our study period 2012-2021. Finally, the Education Register was established in 1974 but also includes information on education completed before this time.

### Sampling

The study population included all residents in Denmark aged at least 30 years between 2012-2021 and born after 1920 (N = 4,258,789). The lower age limit was determined based on the assumption that most people have reached their final educational level by age 30. Moreover, persons born before 1920 were excluded due to limited information on education.

Eligible individuals were followed from the day they reached the inclusion age or year and until time of diagnosis, death, emigration or the end of the study period, whichever came first. Our sample included all 833 single disease analyses. However, individuals who had been registered with a specific disease prior to study entrance were excluded from the analysis of that given disease, but were included in the analysis of other diseases. Moreover, individuals with multiple diagnoses contributed to the analyses of each of their diagnoses separately. Hence, we ignored the order of diagnoses. Information on age, sex, birth cohort and education were determined at the time of inclusion.

### Diseases

Diseases were defined based on the International Classification of Diseases, 10^th^ revision (ICD-10), which classifies diseases with six level codes, which adds up to more than 69,000 unique diagnostic codes. We aggregated diagnostic codes on the third level, which condensed the information to a total of 1,591 diseases. Due to the lower age limit of the study population, we selected only diseases that mostly occur after the age of 30 years. Thus, conditions with a least 25% of cases occurring before the age of 30 years were excluded. Furthermore, we excluded diagnostic codes reported less than 100 times during our study period for statistical power. In the end, a total of 833 diagnostic codes were studied.

Time of diagnosis was defined as the time of the first contact with a hospital where an ICD-10 code was registered as either diagnosis type A (action diagnosis) or B (other diagnosis).

### Education

Educational level was defined according to the International Standard Classification of Education (ISCED) version 2011 and further categorized into three levels: “low”, “medium” and “high” (UNESCO Institute for Statistics, 2012). Low educational level included early childhood education, primary education, lower secondary education and higher secondary education. Medium educational level included post-secondary non-tertiary education such as high school and vocational training. High educational level included tertiary education such as Bachelor’s and Master’s studies.

### Analysis

We estimated age-standardized incidence rates for each combination of educational level and diagnostic code. A Poisson regression model was used for this purpose and allowed us to further adjust for sex and birth cohort. Based on the regression model, age specific incidence rates were estimated for five-year intervals and standardized using the weights from the European Standard Population 2013 to enable a direct comparison across educational groups (Eurostat Task force, 2013). The age-standardized incidence rates were then calculated as the sum of the weighted age-specific incidence rates with medium education as the reference group.

Additionally, we performed complementary analyses using the same approach as described above, however, focusing on noncommunicable (NCDs) and communicable (CDs) diseases respectively, as defined by the Global Burden of Disease study (Mathers et al., 2006). According to these defined disease groups, NCDs accounted for 58.83% and CDs 2.59% of the disease burden.

## Results

Our sample included a total of 4,258,789 individuals of which 26% had an educational attainment categorized as low, 40% had completed a medium length education and 34% had completed a higher education at the time of inclusion. Table 1 presents the distribution of population characteristics. The composition of the population in each of the three groups differed in terms of sex as the majority of the lower (54%) and higher (55%) educated populations were female, while males constituted the majority of the medium educated population (55%). With 38% of the lower educated being at least 65 years of age, compared to 20% among the medium education and 12% among the higher educated, the populations differed substantially in age distribution. Simultaneously, the lower educated population consisted of more individuals from older birth cohorts than the medium and higher educated. This reflects the secular trend of educational expansion, which results in a marked decrease in lower education as well as a marked increase in higher education over time and within younger cohorts.

**Table 1.** Population characteristics N=4,258,789

|  | **Low education**<br>**N=1,094,906**<br>**(25.7%)** | **Medium education**<br>**N=1,722,668**<br>**(40.4%)** | **High education**<br>**N=1,444,247**<br>**(33.9%)** |
| --- | --- | --- | --- |
| **Sex** |  |  |  |
| Female | 586,403 (53.6%) | 779,434 (45.3%) | 785,876 (54.5%) |
| Male | 508,017 (46.4%) | 942,368 (54.7%) | 656,691 (45.5%) |
| **Age** |  |  |  |
| 30-34 years | 161,408 (14.7%) | 357,279 (20.8%) | 477,056 (33.1%) |
| 35-39 years | 67,518 (6.2%) | 169,073 (9.8%) | 176,105 (12.2%) |
| 40-44 years | 76,460 (7.0%) | 182,184 (10.6%) | 153,407 (10.6%) |
| 45-49 years | 88,790 (8.1%) | 205,473 (11.9%) | 139,930 (9.7%) |
| 50-54 years | 91,767 (8.4%) | 162,859 (9.5%) | 115,978 (8.0%) |
| 55-59 years | 98,910 (9.0%) | 148,390 (8.6%) | 108,588 (7.5%) |
| 60-64 years | 95,546 (8.7%) | 157,719 (9.2%) | 98,148 (6.8%) |
| 65-69 years | 115,286 (10.5%) | 142,952 (8.3%) | 78,797 (5.5%) |
| 70-74 years | 98,408 (9.0%) | 85,673 (5.0%) | 42,748 (3.0%) |
| 75-79 years | 81,850 (7.5%) | 55,942 (3.2%) | 26,012 (1.8%) |
| 80-84 years | 65,875 (6.0%) | 33,553 (1.9%) | 15,872 (1.1%) |
| 85-89 years | 45,530 (4.2%) | 18,036 (1.0%) | 8,650 (0.6%) |
| 90+ years | 7,072 (0.6%) | 2,669 (0.2%) | 1,276 (0.1%) |
| **Birth cohort** |  |  |  |
| 1920-1929 | 90,448 (8.3%) | 38,215 (2.2%) | 18,437 (1.3%) |
| 1930-1939 | 166,935 (15.3%) | 118,948 (6.9%) | 56,605 (3.9%) |
| 1940-1949 | 217,148 (19.8%) | 275,914 (16.0%) | 154,457 (10.7%) |
| 1950-1959 | 187,581 (17.1%) | 303,577 (17.6%) | 213,460 (14.8%) |
| 1960-1969 | 170,952 (15.6%) | 384,246 (22.3%) | 270,335 (18.7%) |
| 1970-1979 | 132,191 (12.1%) | 324,473 (18.8%) | 329,036 (22.8%) |
| 1980-1990 | 129,165 (11.8%) | 276,429 (16.1%) | 400,237 (27.7%) |

### Incidence across all diseases

We found educational differences in incidence rates across all 833 diseases examined. Results for all diagnoses codes can be viewed here: https://annaverajp.shinyapps.io/incidence_atlas/. For 569 diseases, we observed a negative educational gradient in which the incidence rates decreased with a higher educational level. While for 77 diseases, we observed an inverse educational gradient. Among the remaining diagnoses, the medium educated had the lowest incidence rate for 103 diseases and the highest for 84 diseases.

Differences in incidence rates were not limited to a few particular disease areas but was observed across all the 16 ICD-10 chapters studied as shown in Figure 1. Each disease is represented by a red and blue dot in the figure. Each of which reflect a mortality rate ratio between low (red dot) or high (blue dot) educational level and medium educational level. These mortality rate ratios are plotted on a log scale on the x-axis and placed on the y-axis according to their respective ICD-10 chapter. The vertical dashed line from log(mortality rate ratio) = 0 shows whether the dots indicate a lower (left side of the line) or higher (right side of the line) mortality rate compared to the reference group (medium educated). The general pattern observed is that incidence rates are lowest among the higher educated and highest among the lower educated. This pattern is observed in all chapters studied, however, most apparent in diseases of the blood and blood-forming organs (chap. 3); mental and behavioral disorders (chap. 5); diseases of the circulatory system (chap. 9); diseases of the digestive system (chap. 11); and symptoms, signs and abnormal clinical and laboratory findings (chap. 18). For other chapters, the pattern is more mixed as for instance neoplasms (chap. 2); diseases of the nervous system (chap. 6); diseases of the eye and adnexa (chap. 7); diseases of the ear and mastoid process (chap. 8); and diseases of the skin and subcutaneous tissue (chap. 12).

**Figure 1.**
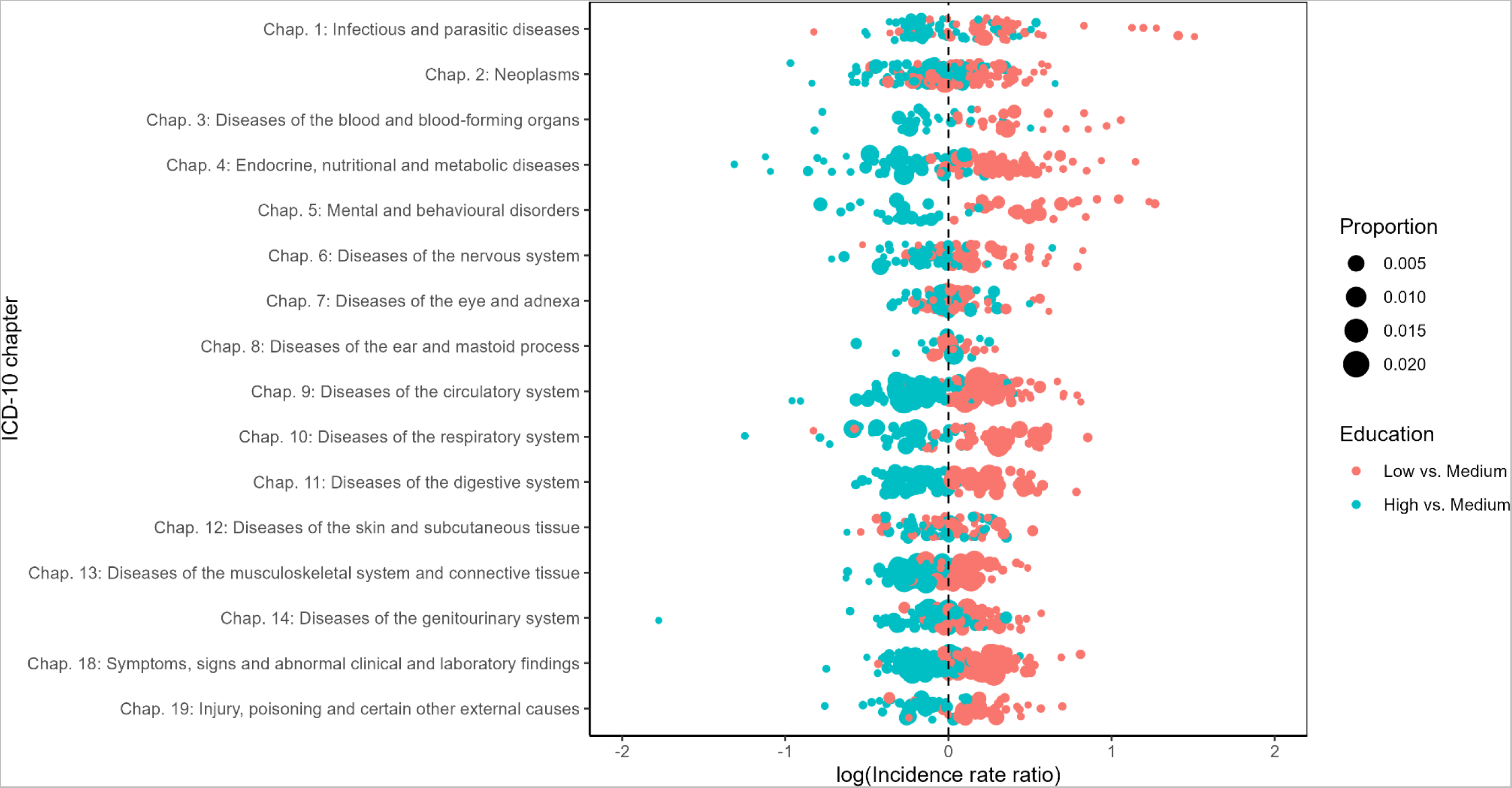
Age-standardized incidence rate ratios dependent on educational achievements.

As Figure 1 shows, the general pattern of educational inequalities across ICD-10 chapters, Figure 2 zooms in on neoplasms (chap. 2), to provide more detail on educational inequalities within ICD-10 chapters. Similar plots of the other chapters can be viewed here: https://annaverajp.shinyapps.io/incidence_atlas/. Across the neoplasms, the vast majority of associations displayed a negative educational gradient – the higher the education the lower the incidence. Nonetheless, few showed a positive gradient. This was for example to case for neoplasms of the breast (C50, D05, D24); skin (C43-44, D03-04, D22-23); prostate (C61); eye, brain and other parts of central nervous system (C69-71, D31-33); and the thyroid gland (C73, D34).

**Figure 2:**
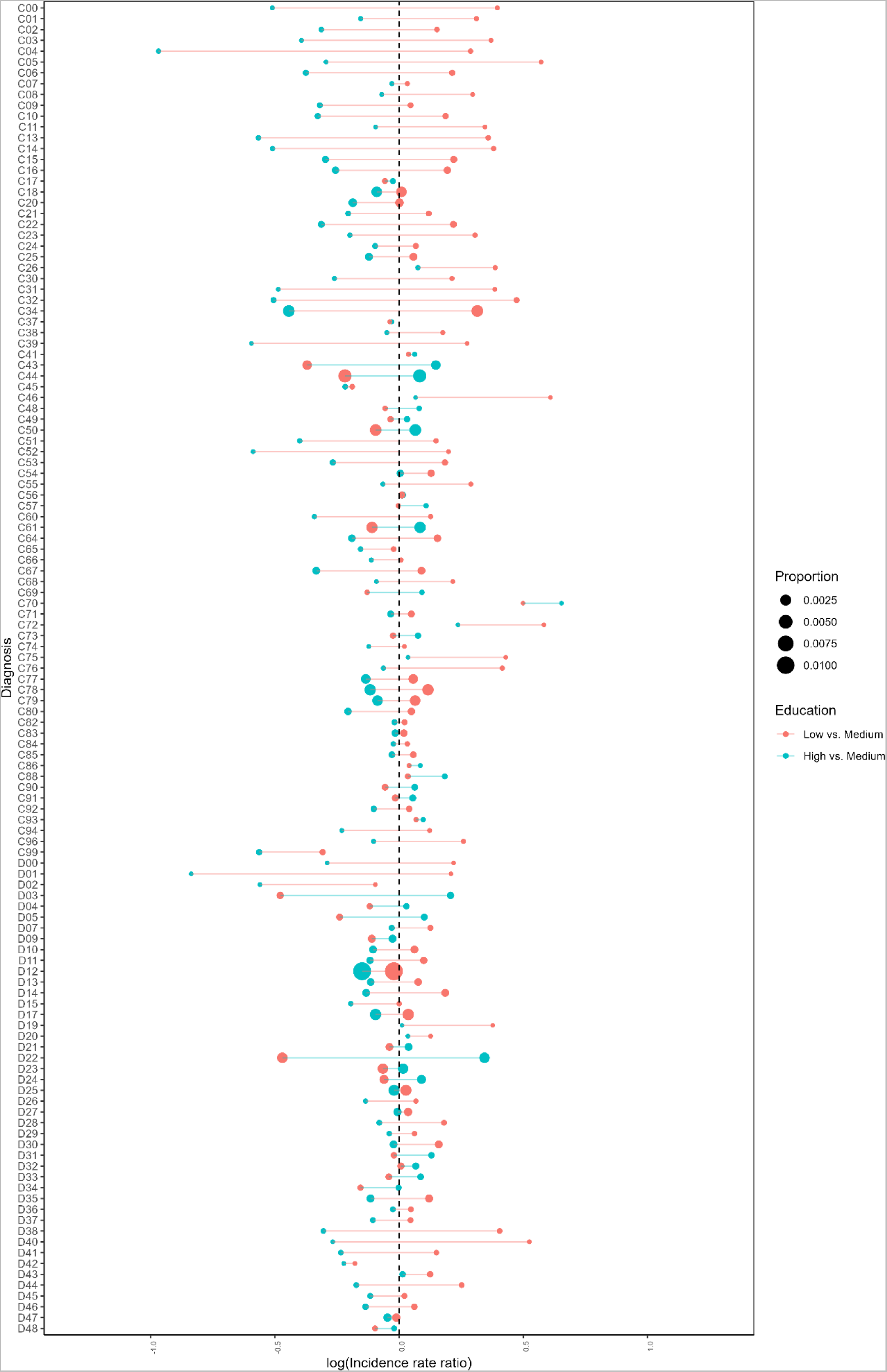
Age-standardized incidence rate ratios for neoplasms (ICD-10 chapter 2) dependent on educational achievements.

### Educational inequalities in the incidence of noncommunicable diseases (GBD)

When we examined non-communicable diseases using disease categories as defined by the Global Burden of Disease study, we found a negative educational gradient for most disease categories (Figure 3). For diabetes, endocrine disorders, cardiovascular diseases, respiratory diseases, digestive diseases, skin diseases, musculoskeletal diseases and oral conditions, the lower educated always had the highest incidence rate and most often the higher educated had the lowest. Among the malignant neoplasms only cancer of the breast, skin and prostate showed a positive gradient which is in line with results from our atlas of ICD-10 diagnostics codes (Figure 2). Regarding non-specified ‘other’ neoplasms, medium educated residents had a slightly higher incidence rate than both the lower and higher educated. The majority of the neuropsychiatric conditions had a negative educational gradient, however, the higher educated had the highest incidence rates for Parkinson’s disease and insomnia, while the lower educated had an incidence rate very similar to the medium educated. The same were the case for hearing loss and other sense organ disorders among sense organ diseases and benign prostatic hypertrophy among the genitourinary diseases.

**Figure 3.**
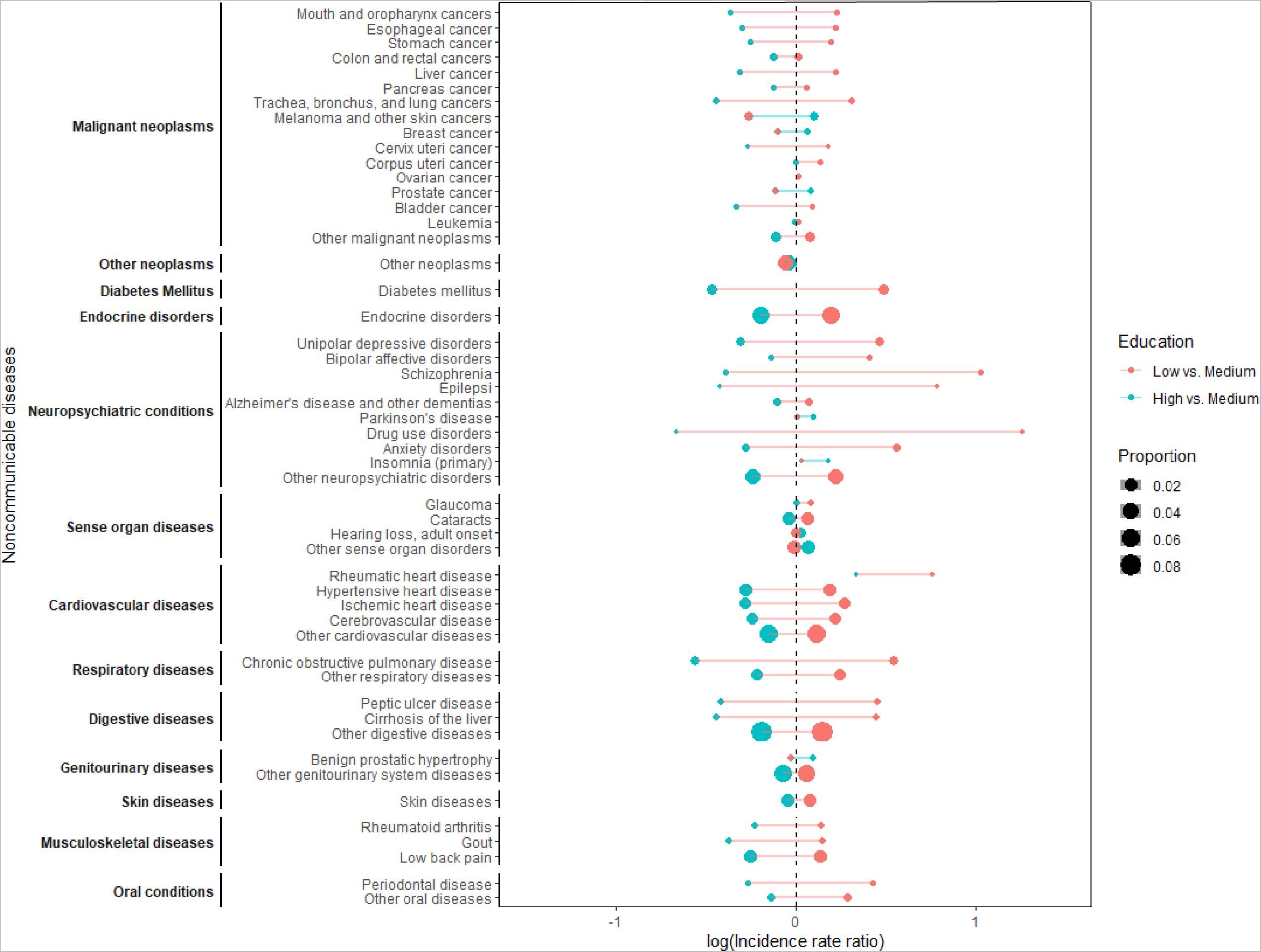
Age-standardized incidence rate ratios for non-communicable diseases defined by the Global Burden of Disease study dependent on educational achievements.

**Figure 4:**
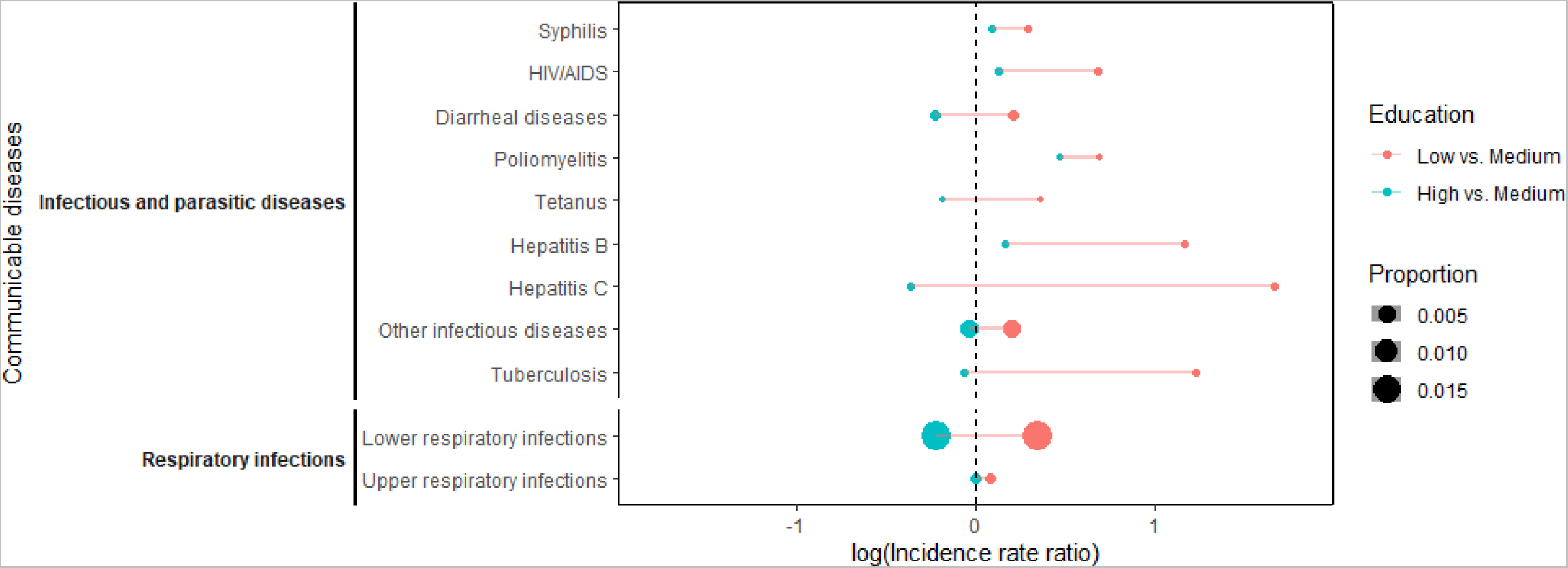
Age-standardized incidence rate ratios for communicable diseases defined by the Global Burden of Disease study dependent on educational achievements.

### Educational inequalities in the incidence of communicable diseases (GBD)

Examining communicable diseases using disease categories from the Global Burden of Disease Study, showed that lower educated had the highest incidence rates for all the CDs studied. Medium educated had the lowest incidence rates for syphilis, HIV/AIDS, poliomyelitis and Hepatitis B, while higher educated had the lowest for the remaining CDs studied.

## Discussion

We document a socioeconomic gradient for a lion’s share of the universe of diseases. This highlights that the educational gradient in health is not confined to the conditions studied most frequently but that it is a phenomenon that affects nearly all disease categories. Educational difference exists for most diseases and primarily such that those with the shortest educations have the highest incidence. This in and of itself is remarkable since the diseases studied are heterogeneous in terms of etiology and diagnosis. Our findings also show another type of heterogeneity: The magnitude of the educational inequalities within ICD-10 chapters can vary substantially. This reflects the heterogeneity in the causal processes that are responsible for linking educational attainment with the incidence and diagnosis of diseases.

This study was made possible by the Danish registries which provide a unique opportunity to examine associations between education and disease incidence in detail for the entire population. This provides the numbers needed for the phenome-wide approach. It also eliminates the selection bias present in studies based on insurance claims data.

The population-wide data used in this study also have limitations. First, the data comes from hospital contacts (Schmidt et al., 2015). This means that the incidences are conditional on seeking treatment at a hospital. As a result, health seeking behavior and the physician referral patterns will affect the incidences. There is good reason to believe that the educational attainment of patients exerts an influence on this: Well-educated citizens may have better de facto access to services so that even though the access is free and fair, the distribution of services consumed do not reflect the disease burden one-to-one. The quality of the information stored in the DNPR is generally good, but it differs substantially from disease to disease (Schmidt et al., 2015). It is unknown if there are educational differences in the quality of information, but it is not unlikely, which may distort our estimates. Also, diseases that are primarily treated in general practice or by other non-hospital specialists are not recorded in the DNPR and is consequently not included in this present study.

The educational expansion and the resulting difference in educational attainment across cohorts is handled through age standardization, but the results may still be influenced by cohort effects that limits the generalizability of our findings to future cohorts. Our phenome-wide approach is built on scanning across diseases with different debut ages in a population that has reached an age where most will have completed their educational career. This means that the age range at inclusion might make more sense for some diseases than for other. For diseases with an early age of diagnosis many cases may be excluded (e.g. schizophrenia), while for others the incidence is essentially null at the age of inclusion (e.g. dementia). This is important to consider in the interpretation of results. While our set of outcomes is comprehensive, we still omit a large number of diseases from our analysis to minimize the number of false positives. This does not mean that no educational differences could be observed in those groups, only that we did not examine it.

For previously studied diseases our results are congruent with the literature. This is true for cardiovascular diseases and for cancer, which has been studied comprehensively (Christensen et al, 2016; Dalton et al., 2008; Diderichsen et al., 2012). Direct comparison is, however, often hampered by differences in population, case definition, analysis and measures of association. For some diseases the literature is sparse or non-existing. Often, the causal pathways from education to incident disease is less clear for comparatively understudied diseases. For example, a UK study from 2016 reported an inverse association between socioeconomic position and age-related macular degeneration in a population survey (Yip et al., 2015). Some of this may be due to educational differences in exposure to causal agents, but may also be influenced by educational differences in health care seeking behavior (Zhang et al., 2013). Moreover, comparability between studies is affected by differences in their context. We conducted our study in Denmark - a highly egalitarian country. Differences in income are reduced through a progressive tax system which finances free education with universal access. Hence, socioeconomic inequalities are reduced. Despite the high level of egalitarianism, we do find educational inequalities in most diseases studied. They might, however, be more pronounced in less egalitarian countries. We focused on educational inequalities in disease incidence but our analysis could be extended to other measures of socioeconomic position such as income or occupation. Extending the atlas with multiple socioeconomic characteristics which may be differently associated with disease incidence would provide a rich understanding of socioeconomic inequalities across a very wide selection of diseases. Why are there so striking similarities in the educational pattern in disease incidence? Shared physiology is central as many diseases are caused by the malfunction of an organ system, which in turn can manifest itself in different diseases (Amell et al., 2018). This type of horizontal pleiotropy is an important driver of similarity. Dyslipidemia, for example, can lead to several different cardiovascular diseases (The Emerging Risk Factors Collaboration, 2012). The pleiotropy can also be vertical: Poorly regulated diabetes mellitus or depression may have down-stream effects on the risk of other diseases (Knight et al., 2017; Penninx et al., 2013). Beyond the biological processes within the human body, there are also causes that act in the physical and social environment. Tobacco smoking is the most extensively documented cause of disease in existence (Musk and De Klerk, 2003) and has a strong educational gradient (Palipudi et al., 2012). The same holds true for many other causes of ill health: Environmental pollutants, work exposures, poor nutrition, lack of access to physical activity etc. (Olsen et al., 2019; WHO, 2010; Monden, 2005). Jointly, these shared mechanisms will work to cause educational gradients across diseases.

In conclusion, we believe that our atlas provides a full and detailed representation of the association between education and disease occurrence. It brings attention to all diseases – not only the most prevalent – and makes it possible to examine the role of education across the universe of diseases.

## Data Availability

It is possible for individual researchers to gain access to the relevant microdata through Statistics Denmarks' research services and reconstruct the data used in this paper.

## Acknowledgements

We would like to express our sincere gratitude to Rudi G. J. Westendorp for his invaluable guidance, important input and for challenging us in numerous discussions throughout the research process. His contributions and support have helped us enormously in shaping the direction of the study. We also wish to thank Jim Vaupel for valuable discussions in the very early stages of the project.

